# Polygenic modulation of lipoprotein(a)-associated cardiovascular risk

**DOI:** 10.1101/2020.02.22.20026757

**Authors:** Mark Trinder, Liam R. Brunham

## Abstract

**Aims:** Elevated levels of lipoprotein(a) are one of the strongest inherited risk factors for coronary artery disease (CAD). However, there is variability in cardiovascular risk among individuals with elevated lipoprotein(a). The sources of this variability are incompletely understood. We assessed the effects of a genomic risk score (GRS) for CAD on risk of myocardial infarction among individuals with elevated lipoprotein(a).

**Methods:** We calculated CAD GRSs for 408,896 individuals of British white ancestry from the UK Biobank using 6.27 million common genetic variants. Lipoprotein(a) levels were measured in 310,020 individuals. The prevalence and risk of myocardial infarction versus CAD GRS percentiles were compared for individuals with and without elevated lipoprotein(a) defined as ≥120 or 168 nmol/L (≈50 or 70 mg/dL, respectively).

**Results:** Individuals with elevated lipoprotein(a) displayed significantly greater CAD GRSs than individuals without elevated lipoprotein(a), which was largely dependent on the influence of genetic variants within or near the *LPA* gene. Continuous levels of CAD GRS percentile were significantly associated with risk of myocardial infarction for individuals with elevated lipoprotein(a). Notably, the risk of myocardial infarction for males with elevated lipoprotein(a) levels, but a CAD GRS percentile in the lower quintile (<20^th^ percentile), was less than the overall risk of myocardial infarction for males with non-elevated lipoprotein(a) levels (hazard ratio [95% CI]: 0.79 [0.64-0.97], p=0.02). Similar results were observed for females.

**Conclusion:** These data suggest that CAD genomic scores influence cardiovascular risk among individuals with elevated lipoprotein(a) and may aid in identifying candidates for preventive therapies.

## INTRODUCTION

Lipoprotein(a) is a plasma lipoprotein composed of a low-density lipoprotein particle that is covalently linked to apolipoprotein(a) by a disulfide bond. Lipoprotein(a) levels are among the strongest inherited risk factors for cardiovascular disease^1–9^, and Mendelian randomization studies suggest that elevated levels of lipoprotein(a) are causal for aortic stenosis^10^, heart failure^11^, ischemic stroke^12^, and coronary artery disease (CAD)^1,13^. Plasma levels of lipoprotein(a) are largely determined by genetic factors which include single-nucleotide variants and copy number variants in the kringle IV type 2 (KIV-2) domain of the *LPA* gene^5,6,14,15^.

Elevated lipoprotein(a) is a common condition that is estimated to affect 1 in 5 individuals of European ancestry (plasma lipoprotein(a) levels greater than 50 mg/dL)^16^. Although elevated lipoprotein(a) is associated with increased cardiovascular risk, the optimal treatment of elevated lipoprotein(a) is uncertain. While there are currently no approved therapies for lowering lipoprotein(a), antisense oligonucleotides that target apolipoprotein(a) and reduce lipoprotein(a) levels by up to 80% have been developed and are currently being studied in clinical trials^17–19^. At the individual level, there is substantial variability in cardiovascular risk among individuals with elevated lipoprotein(a)^1,20,21^, and it remains unclear whether isolated elevated lipoprotein(a) in the absence of other cardiovascular risk factors merits treatment to reduce cardiovascular risk.

Recent work suggests that genomic risk scores (GRSs), which summarize the association of millions of common genetic variants with risk of CAD into a single value, may be a useful tool for improving risk prediction of CAD^7,22–25^. Here we tested the hypothesis that, among individuals with elevated lipoprotein(a), the risk of CAD would be modulated by a GRS for CAD, and that this may be useful for risk stratification.

## METHODS

### Study population

We studied participants from the UK Biobank, a large population-based prospective cohort study from the United Kingdom that aims to improve the prevention, diagnosis and treatment of disease by following the health and well-being of approximately 500 000 individuals. Individuals were enrolled between 2006-2010 and were between 40-69 years-of-age. Individuals underwent deep phenotyping at study enrollment and DNA was collecting for genotyping, as previously described^26^. The UK Biobank resource was approved by the UK Biobank Research Ethics Committee, and all participants provided written informed consent to participate in the study. This study was approved by the UK Biobank (application ID: 42857) and by the Clinical Research Ethics Board of the University of British Columbia (H18-02181).

### Biochemical measurements

Biochemical measurements were assessed at the time of study enrollment (Supplemental Methods; Supplemental Table 1). Lipoprotein(a) was measured using an immuno-turbidimetric method on the Beckman Coulter AU5800 platform (Randox Bioscience, UK), which is essentially isoform insensitive^27^. Where indicated, lipoprotein(a) concentration was converted from nmol/L to mg/dL by dividing values by 2.4^3,28^. Elevated lipoprotein(a) was defined as lipoprotein(a) levels ≥ 120 nmol/L (≈50 mg/dL) or ≥ 168 nmol/L (≈70 mg/dL)^27,29,30^. For individuals taking cholesterol-lowering medication, total cholesterol and low-density lipoprotein cholesterol levels were adjusted by multiplying on-treatment lipid levels by 1.43, corresponding to an estimated 30% reduction in low-density lipoprotein cholesterol^31,32^.

**Table 1.** Enrollment characteristics and cardiovascular history of the study group. High-density lipoprotein cholesterol (HDL-C), interquartile range (IQR), low-density lipoprotein cholesterol (LDL-C) standard deviation (SD).

| Characteristic | Measurement | Values |
| --- | --- | --- |
| n | no. | 408896 |
| Age | mean (SD) | 56.9 (8.0) |
| Female sex | no. (%) | 221082 (54.1) |
| Hypertension | no. (%) / n | 110888 (27.2)<br>/ 408256 |
| Severe hypercholesterolemia | no. (%) / n | 31978 (8.2)<br>/ 389158 |
| Diabetes mellitus | no. (%) / n | 19773 (4.8)<br>/ 408003 |
| Obesity | no. (%) / n | 98879 (24.3)<br>/ 407600 |
| Current smoker | no. (%) / n | 41320 (10.1)<br>/ 407461 |
| Total cholesterol (mmol/L) | median (IQR) / n | 5.8 (1.55)<br>/ 389875 |
| LDL-C (mmol/L) | median (IQR) / n | 3.61 (1.16)<br>/ 389158 |
| Triglycerides (mmol/L) | median (IQR) / n | 1.50 (1.11)<br>/ 389567 |
| HDL-C (mmol/L) | median (IQR) / n | 1.40 (0.50)<br>/ 356840 |
| Lipoprotein(a) (nmol/L) | median (IQR) / n | 20.1 (50.88)<br>/ 310020 |
| Hemoglobin A1c (mmol/mol) | median (IQR) / n | 35.2 (5.1)<br>/ 389771 |
| C-reactive protein (mg/L) | median (IQR) / n | 1.33 (2.1)<br>/ 389036 |

### Calculation of coronary artery disease genomic risk scores

The weightings used to calculate CAD GRSs were obtained from the Cardiovascular Disease Initiative Knowledge Portal and can be accessed at www.broadcvdi.org^7,22^. CAD GRSs were calculated for individuals of British white ancestry using imputed genetic data that were not flagged as outliers for excess missingness or heterozygosity^26^. CAD GRSs were calculated using 6.27 million of the 6.63 million potential single-nucleotide variants described by Khera et al. (2018). Variants that displayed deviation from Hardy-Weinberg equilibrium (p<1×10^−6^), a genotyping rate <95%, or minor allele frequency <1% were removed. The PLINK score function was used to multiply the number of alleles associated with adjusted β-coefficient for increased risk of CAD at each single-nucleotide variant by its respective linkage disequilibrium-adjusted weight and sums these products across all available variants to generate a GRS for each individual^23,33^. CAD GRSs were also calculated after excluding genetic variants located within 7.5 million base pairs upstream and downstream of the *LPA* gene. CAD GRSs were converted into percentiles relative to the distribution of CAD GRSs in the UK Biobank study population.

The top 100 genetic variants used in the calculation of CAD GRSs were manually curated for association with known cardiovascular risk factors using the NHGRI-EBI Genome-Wide Association Study Catalog^34^.

### Definition of cardiovascular events

Phenotypes, including the primary outcome of myocardial infarction, were defined using reports from medical history interviews occurring at enrollment, International Classification of Diseases (ICD)-9^th^ and −10^th^ Revision codes, and death registry records (Supplemental Methods, Supplemental Table 2). Events occurring before and after enrolment were included unless otherwise stated. Events occurring prior to enrolment were identified by either self-reported medical history and/or previous hospital admission within an electronic health record. Incident events were defined by hospital admission with an electronic health record entry or death records. Events were censored on the date of loss-to-follow-up or if individuals remained event-free up to March 31, 2017.

### Statistical analyses

All statistical analyses were performed using R version 3.6.0 software (R Core Team, 2019). Individuals with missing values were excluded from analyses.

Chi-square tests were used for contingency analyses. For comparison of 2 groups, data were analyzed with an unpaired t-test or Mann–Whitney U test as appropriate. For comparison of more than 2 groups, data were analyzed with one-way analysis of variance test (with Tukey’s multiple comparison post hoc tests) or Kruskal-Wallis test, as appropriate (with Dunn’s multiple comparison post hoc tests).

The distributions of CAD GRS percentiles were compared between individuals with traditional cardiovascular risk factors determined at study enrollment that included: hypertension, severe hypercholesterolemia (low-density lipoprotein cholesterol levels ≥ 4.9 mmol/L), diabetes mellitus, obesity (body mass index ≥ 30 kg/m^2^), current smoker status, insufficient physical activity (classification of low activity level according to the International Physical Activity Questionnaire), insufficient fruit and vegetable intake (< 3 servings per day; sum of cooked vegetable intake, fresh fruit intake, and salad/raw vegetable intake), and strata of lipoprotein(a) levels (<72, 72-120, 120-168, and ≥168 nmol/L).

The prevalence of cardiovascular events of interest were calculated for each decile of CAD GRS percentile. Linear regression models were used to assess the correlation between cardiovascular disease prevalence and CAD GRS percentile using elevated lipoprotein(a) status as a covariate or interactive term

Time-to-event analyses were analyzed with the “survival” version 2.43-3 package for R with Log-rank tests. Hazard ratios (HRs) and 95% confidence intervals (CIs) were calculated using Cox regression models stratified by sex with age of event as a time scale. All Cox regression models were adjusted for the first 4 principal components of ancestry and genotyping array.

Statistical significance was claimed when two-sided p-values were <0.05.

## RESULTS

### Characteristics of individuals from the UK Biobank

This study included 408,896 individuals of British white ancestry from the UK Biobank study, of whom 310,020 had lipoprotein(a) levels measured at enrollment (Table 1). The average age of enrollment was 56.9 years-of-age (standard deviation: 8.0 years) and 54.1% were female.

### Genomic risk scores for coronary artery disease associate with the prevalence of traditional cardiovascular risk factors

First, we assessed how CAD GRSs were associated with traditional cardiovascular risk factors. Individuals with hypertension, severe hypercholesterolemia, diabetes mellitus, obesity, and a current smoker status displayed significantly higher CAD GRS percentiles than those without the cardiovascular risk factor of interest (Mann-Whitney U test: p=0.0006 for smoking status, p<0.0001 for others; Figure 1a-e; Supplemental Table 3). There was also a significant stepwise increase in the median CAD GRS percentile as the number of traditional CAD risk factors per an individual increased (Kruskal-Wallis test: p<0.0001; Figure 1f). Alternatively, CAD GRSs did not display an association with insufficient physical activity (Mann–Whitney U test: p=0.40) or an insufficient daily intake of fruits and vegetables (Mann–Whitney U test: p=0.40) (Supplemental Figure 1).

**Figure 1.**
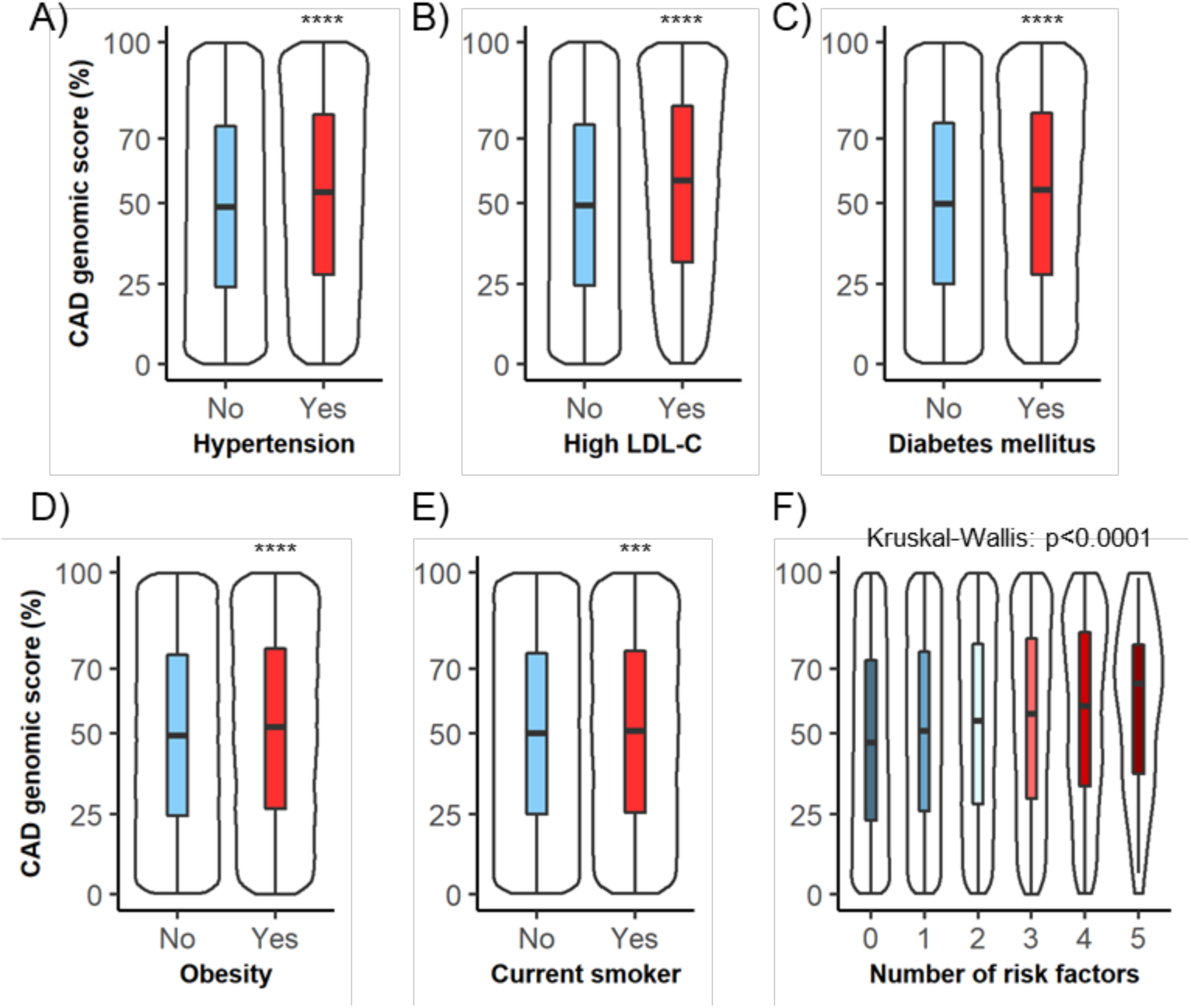
Genomic scores for coronary artery disease associate with traditional cardiovascular risk factors. The distribution of coronary artery disease genomic score percentiles are displayed for traditional cardiovascular risk factors assessed at study enrollment which include: (A) hypertension, (B) high low-density lipoprotein cholesterol (LDL-C ≥ 4.9 mmol/L), (C) diabetes mellitus, (D) obesity (body mass index ≥ 30 kg/m^2^), (E) current smoking status, and (F) a sum of the aforementioned risk factors. Boxplots display the median and interquartile range. The boxplot’s whiskers show the range and the accompanying density distribution. *** p<0.001, **** p<0.0001.

### Individuals with elevated lipoprotein(a) have higher genomic risk scores for coronary artery disease

CAD GRS percentiles displayed a significant and strong association with lipoprotein(a) levels (Kruskal–Wallis test: p<0.0001; Figure 2A). Notably, individuals with lipoprotein(a) levels greater or equal to 120 nmol/L (≈50 mg/dL) displayed a median CAD GRS of 65.2 percentile. Individuals with elevated lipoprotein(a) tended to have CAD GRSs that were comparable to individuals with multiple traditional cardiovascular risk factors at the time of study enrollment (*i*.*e*. the median CAD GRS percentile for individuals with hypertension, severe hypercholesterolemia, diabetes mellitus, obesity, and current smoker status was 65.5 percentile). The higher CAD GRS percentile observed among individuals with elevated lipoprotein(a) was dependent on genetic loci within and around the *LPA* gene. When CAD GRSs were re-calculated after the exclusion of DNA variants located 7.5 million base pairs up- or downstream of the *LPA* gene, the difference in CAD GRSs across strata of lipoprotein(a) levels was attenuated (Kruskal– Wallis test: p=0.02; Figure 2B; Supplemental Table 4).

**Figure 2.**
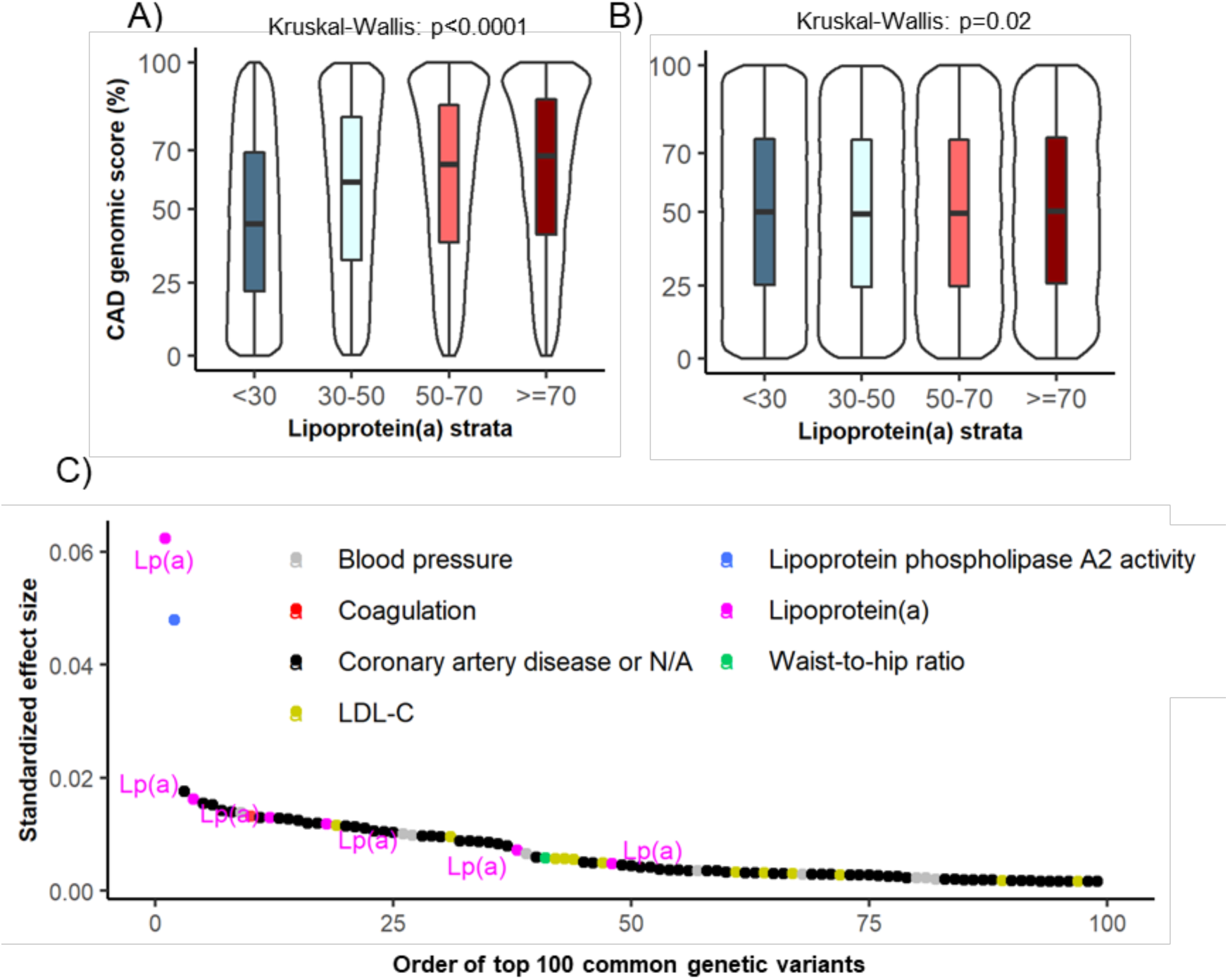
Elevated lipoprotein(a) is a major determinant of coronary artery disease genomic risk scores. The distribution of coronary artery disease genomic risk score percentiles are displayed across strata of lipoprotein(a) levels calculated (A) with and (B) without genetic variants in proximity to the *LPA* gene. Boxplots display the median and interquartile range. The boxplot’s whiskers show the range and the accompanying density distribution. (C) The standardized effect sizes for the top 100 genetic variants used in calculating coronary artery disease genomic risk scores are shown coloured by reported associations with traditional or emerging cardiovascular risk factors in the NHGRI-EBI Genome-Wide Association Study Catalog. Lipoprotein(a) [Lp(a)].

The strong association between CAD GRSs and elevated lipoprotein(a) reflects both the high heritability and atherogenicity of lipoprotein(a) particles. Specifically, the locus in the CAD GRS with the greatest standardized effect size (rs186696265; GRCh37 position 6:161111700:C:T), and hence the greatest influence on CAD GRSs by weight, is known to associate with lipoprotein(a) levels (Figure 2C). Moreover, 6 of the top 100 loci with the largest standardized effect sizes in the CAD GRSs have known associations with lipoprotein(a) levels and include: rs186696265, rs10455872 (6:161010118:A:G). rs55730499 (6:161005610:C:T), rs118039278 (6:160985526:G:A), rs56393506 (6:161089307:C:T), and rs4252185 (6:161123451:T:C) (Figure 2C).

### Association of lipoprotein(a) levels with risk of myocardial infarction

Consistent with previous studies^1,27^, lipoprotein(a) levels were right-skewed among individuals of British white ancestry from the UK Biobank (Figure 3A). The median and interquartile range of lipoprotein(a) levels was 20.1 and 50.9 nmol/L, respectively. The risk of myocardial infarction was significantly associated with lipoprotein(a) levels for both male (adjusted HR [95% CI] 1.06 [1.06-1.07] per 20 nmol/L increase, p<0.0001; Figure 3B) and female UK Biobank participants (adjusted HR [95% CI] 1.03 [1.02-1.05] per 20 nmol/L increase, p<0.0001; Figure 3C).

**Figure 3.**
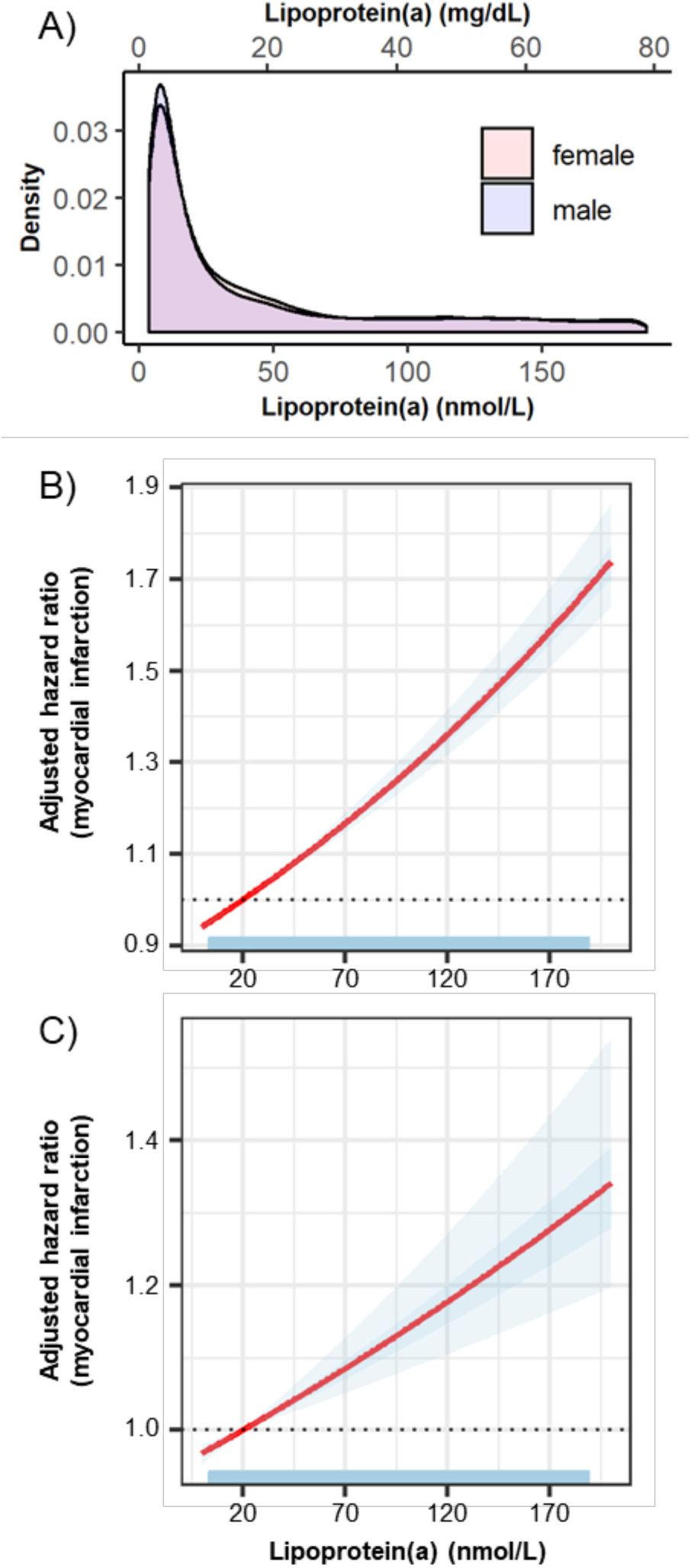
Elevated lipoprotein(a) associates with increased risk of myocardial infarction. (A) The density distribution of lipoprotein(a) levels for UK Biobank participants of British white ancestry is right-skewed. The risk of myocardial infarction versus continuous lipoprotein(a) levels are shown for individuals of (B) male and (C) female sex. Hazard ratios were calculated relative to the median lipoprotein(a) level in this population (20 nmol/L) and adjusted for the first 4 components of genetic ancestry and genotyping array/batch. The darker and lighter blue shading represent the standard error and 95% confidence interval, respectively.

### Risk of myocardial infarction is modulated by coronary artery disease genomic risk score among individuals with elevated lipoprotein(a)

Next, we assessed if CAD GRSs may help explain some of the heterogeneity in risk of CAD that is observed in individuals with elevated lipoprotein(a), defined as ≥120 nmol/L (≈50 mg/dL). The prevalence of myocardial infarction was significantly associated with deciles of CAD GRS percentile in both individuals with and without elevated lipoprotein(a) (Figure 4A). Individuals with elevated lipoprotein(a) displayed a significantly greater prevalence of myocardial infarction relative to individuals with non-elevated lipoprotein(a) levels across all deciles of CAD GRS (β [SE] = 0.786 [0.194], R^2^=0.913, p=0.0009; Figure 4A; Supplemental Table 5). This effect was even more striking when deciles of CAD GRS percentile were calculated after excluding loci associated within and in proximity to the *LPA* gene (β [SE] = 1.494 [0.216], R^2^=0.920, p<0.0001; Figure 4B). Individuals with elevated lipoprotein(a) but a CAD GRS in the first decile had a lower prevalence of myocardial than individuals with non-elevated lipoprotein(a) and a CAD GRS in the tenth decile (2.98% vs 6.61%, Figure 4B). These data indicate that the CAD GRS modulates the risk of myocardial infarction associated with elevated lipoprotein(a). There was no significant interaction between lipoprotein(a) status and CAD GRS that included or excluded loci in and nearby *LPA*.

**Figure 4.**
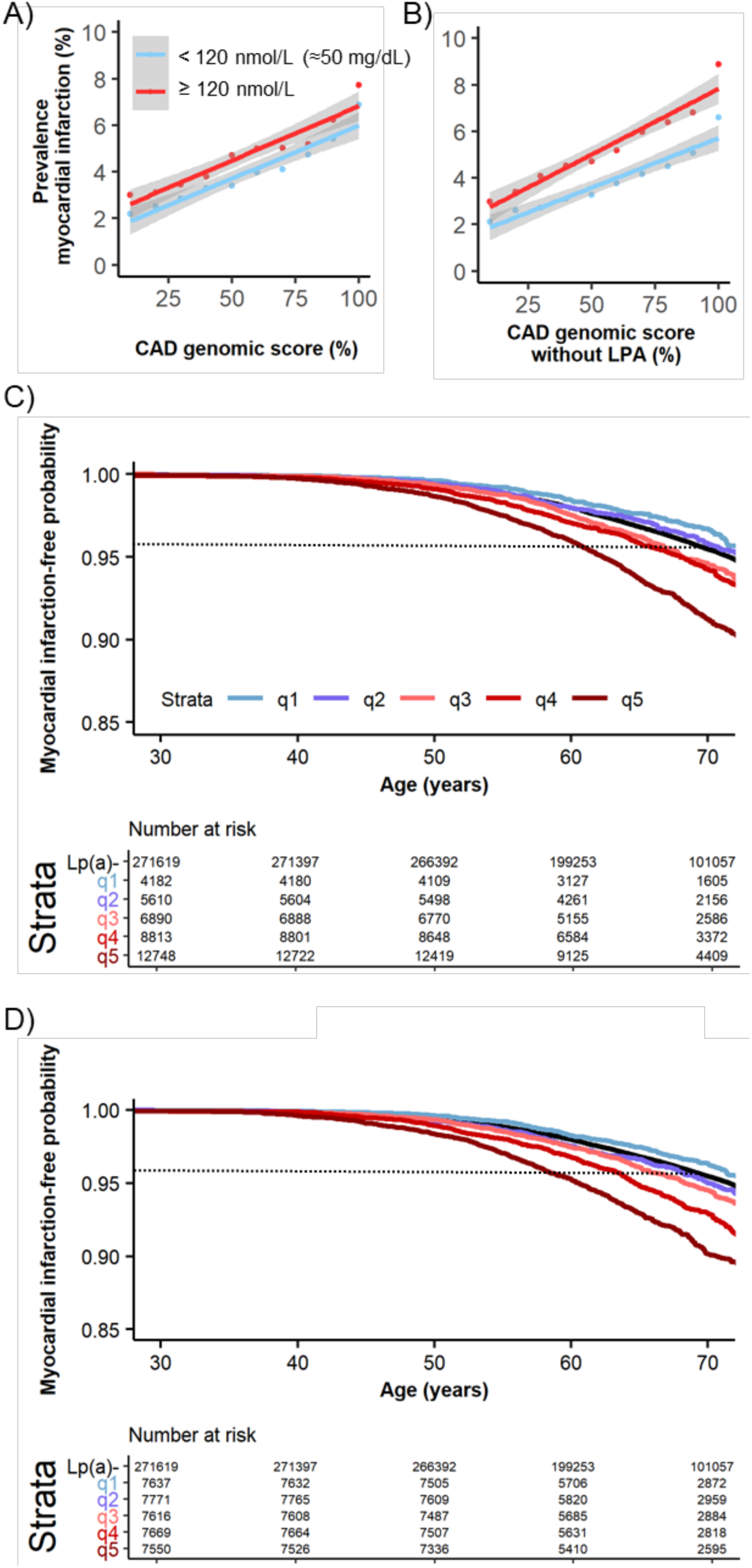
Coronary artery disease genomic risk scores are a modifier of risk of myocardial infarction for individuals with elevated lipoprotein(a). The prevalence of myocardial infarction for each decile of coronary artery disease genomic risk score percentile is shown for scores calculated (A) with and (B) without genetic variants in proximity to the *LPA*. Solid lines depict linear regression analyses and gray shading indicate the associated 95% confidence interval stratified by non-elevated versus elevated lipoprotein(a) levels. Time-to-first-event analyses are displayed for risk of myocardial infarction events stratified by quintiles of coronary artery disease genomic risk score percentile among individuals with elevated lipoprotein(a) (≥ 120 nmol/L) calculated (C) with and (D) without genetic variants in proximity to the *LPA* gene. The solid black line depicts the risk of myocardial infarction among a reference group comprised of all individuals with non-elevated lipoprotein(a) (Lp(a)-, < 120 nmol/L). The horizontal, dotted black line depicts the myocardial infarction-free probability of the non-elevated lipoprotein(a) reference group at 70 years-of-age.

We next examined the cumulative risk of myocardial infarction across time in males and females with elevated lipoprotein(a) as a function of CAD GRS. We observed a significant effect of the CAD GRS on risk of myocardial infarction in these individuals with elevated lipoprotein(a). The adjusted HR for males was 1.28 (95% CI: 1.23-1.33, p<0.0001) and for females was 1.16 (95% CI: 1.08-1.25, p<0.0001) per 20 unit increase in continuous CAD GRS percentile. Notably, the risk of myocardial infarction for individuals with elevated lipoprotein(a) levels, but a CAD GRS percentile in the lower quintile (<20^th^ percentile), was less than or equivalent to the overall risk of myocardial infarction for those with non-elevated lipoprotein(a) levels (adjusted HR [95% CI]: 0.79 [0.64-0.97] and 0.91 [0.66-1.26] with p=0.02 and p=0.58 for males and females, respectively; Figure 4C). Similar results were observed when the risk of myocardial infarction was compared between quintiles of CAD GRS percentiles that excluded loci surrounding the *LPA* gene (adjusted HR [95% CI] were 0.85 [0.73-0.98] and 0.83 [0.64 - 1.07] with p=0.03 and p=0.15 for males and females, respectively; Figure 3D).

Even among individuals with higher lipoprotein(a) levels of ≥168 nmol/L (≈70 mg/dL and comprising the top 3.5 percent of the population distribution), we observed a significant effect of the CAD GRS percentiles on risk of myocardial infarction (Supplemental Figure 2). Specifically, among these individuals, the adjusted HR for myocardial infarction was 1.31 for males (95% CI: 1.22-1.41, p<0.0001) and 1.19 for females (95% CI: 1.04-1.34, p=0.009) per 20 percentile increase in continuous CAD GRS. These findings highlight that genomic background can still modulate the risk of myocardial infarction for individuals with elevated lipoprotein(a) and suggest that the CAD GRS may be useful to stratify cardiovascular risk in individuals identified to have elevated lipoprotein(a).

### Effect of coronary artery disease genomic risk score on other cardiovascular outcomes associated with lipoprotein(a)

Lipoprotein(a) is also likely a causal risk factor for peripheral vascular disease and aortic stenosis^10,35^, and, to a lesser extent, ischemic stroke^12,36^. Since there is considerable overlap of risk factors between these cardiovascular conditions and CAD^37^, we sought to assess if CAD GRSs also modulate the risk of these conditions among individuals with elevated lipoprotein(a). We observed a trend towards increased prevalence of peripheral vascular disease (β [SE]: 0.120 [0.055], p=0.002, R^2^=0.712), aortic stenosis (β [SE]: 0.241 [0.056], p=0.0005, R^2^=0.615), and ischemic stroke (β [SE]: 0.182 [0.072], p=0.02, R^2^=0.361) for individuals with elevated lipoprotein(a), relative to individuals with non-elevated lipoprotein(a) (Supplemental Figure 3; Supplemental Tables 6-8). The strength of the association between prevalence of ischemic stroke and deciles of CAD GRS percentile was weak due to fewer events relative to the other cardiovascular conditions.

## DISUCSSION

Here we report that common genetic factors modify the risk of myocardial infarction and other adverse cardiovascular outcomes among individuals with elevated lipoprotein(a). These data highlight the atherogenicity and high heritably of lipoprotein(a) levels, as reflected by the substantial increases in CAD GRSs observed among individuals with elevated lipoprotein(a). The increased CAD GRSs observed in individuals with elevated lipoprotein(a) were almost entirely explained by single-nucleotide variants near the *LPA* gene having large weighted effect sizes, and thus, a large influence on the CAD GRS.

This work adds to the growing literature highlighting the role that polygenic factors play in modifying the penetrance or expressivity of monogenic conditions, such as familial hypercholesterolemia^22,38–40^. To our knowledge, this is the first study to describe the polygenic modulation of myocardial infarction and other adverse cardiovascular outcomes by CAD GRSs among individuals with elevated lipoprotein(a).

Elevated lipoprotein(a) is a common monogenic condition resulting from genetic variation in and around the *LPA* gene, which includes single-nucleotide variants and copy number variation in the KIV-2 DNA sequence. Variation in the number of KIV-2 repeats results in plasma lipoprotein(a) particles that display considerable interindividual variation in apolipoprotein(a) isoform size (ranging from 300-800 kDa). The number of KIV-2 repeats are also inversely associated with lipoprotein(a) levels. However, lipoprotein(a) levels (*i*.*e*. molar concentration), rather than apo(a) isoform size, appears to be most strongly associated with risk of CAD20,41,42.

Approximately 1 in 5 individuals of European ancestry have lipoprotein(a) levels greater than 120 nmol/L (≈50 mg/mL), which is a common cut-off for estimation of cardiovascular risk and consideration of therapy^30^. Lipoprotein(a) levels greater than 120 nmol/L are also common among individuals of African (1 in 4), South and South East Asian (∼1 in 10), Arab (∼1 in 10), and Latin American (∼1 in 7) ethnicity and also associated with risk of CAD in these populations^20,43,44^. However, the optimal management of patients with elevated lipoprotein(a) is uncertain. Antisense inhibitors that potently lower lipoprotein(a) are currently being studied in clinical trials in subjects with elevated lipoprotein(a) and established cardiovascular disease^17–19^. Our study suggests that the CAD GRS may be useful for identifying high risk patients that are likely to benefit most from lipoprotein(a)-lowering therapy, or other therapies that reduce cardiovascular risk among individuals with elevated lipoprotein(a). In support of this hypothesis, previous studies have shown that individuals with high genetic risk for CAD demonstrate greater absolute risk reduction of cardiovascular events from statins^24,45^ or PCSK9 inhibitors^46^ relative to individuals at lower genetic risk. In addition, the data presented here suggest that a GRS may be useful to de-risk patients with isolated elevated lipoprotein(a), as we observed that a low GRS in such individuals was associated with a risk of myocardial infarction that was similar to, or lower than, that of individuals with non-elevated lipoprotein(a) and a median GRS. The extent to which CAD GRSs can improve risk prediction beyond established clinical risk scores requires further investigation^47^. However, a major advantage of assessing germline genetic variation for cardiovascular risk assessment is that it only needs to be measured once and can be measured early in life, allowing for earlier intervention before other clinical risk factors may emerge^7,25,46^.

This study has some notable strengths and limitations worthy of consideration. Firstly, we were able to use a very large population of more than 300,000 individuals with standardized lipoprotein(a) measurements using an isoform insensitive assay, and genome-wide genotype data. As such, this represents one of the largest population-based studies of lipoprotein(a) to date. Limitations are that the population studied was comprised of individuals of European ancestry and may not be representative of other global populations. While this aspect of the study reduces the risk of bias or confounding due to population stratification, it also limits the generalization of these results to other ancestral populations. Future studies using large populations of individuals from diverse ancestral backgrounds are therefore essential. Indeed, as the calculation and calibration of the CAD GRSs depend on linkage disequilibrium^7,23^, which varies substantially across ancestries, future studies will be needed to derive and validate CAD GRSs in other ancestral groups. The UK Biobank population also displays a healthy volunteer selection bias relative to the overall health and well-being of individuals living across the United Kingdom^48^. Furthermore, the CAD GRS was constructed to best identify individuals at risk of CAD and is likely not as sensitive at identifying individuals at risk of the other cardiovascular conditions described, which include peripheral vascular disease, aortic stenosis, and ischemic stroke. GRSs calibrated specifically for these conditions will likely improve risk prediction.

In summary, genetic factors were shown to significantly modulate the risk of CAD in individuals with elevated lipoprotein(a). Assessment of background polygenic factors may help to personalize risk assessment in individuals with elevated lipoprotein(a) and identify candidates for preventative therapy.

## Data Availability

Data are available from UK Biobank.

## ACKNOWLEDGEMENTS

We acknowledge the participants and individuals involved in the UK Biobank studies; the developers of software tools used in this study; and Compute Canada for establishing computational resources and providing support.

